# Characteristics and healthcare use of patients attending virtual walk-in clinics: a cross-sectional analysis

**DOI:** 10.1101/2022.02.28.22271640

**Authors:** Lauren Lapointe-Shaw, Christine Salahub, R. Sacha Bhatia, Laura Desveaux, Richard H. Glazier, Lindsay Hedden, Noah M. Ivers, Danielle Martin, Sheryl Spithoff, Yingbo Na, Mina Tadrous, Tara Kiran

## Abstract

**Importance:** Virtual walk-in clinics have proliferated since the onset of COVID-19. Yet, little is known about those who participate in this care model, and how virtual walk-in clinics contribute to care continuity and patient healthcare utilization.

**Objectives:** To describe the characteristics and healthcare use of patients using virtual walk-in clinics compared to the general population, and a subset that received any virtual family physician visit.

**Design:** This was a retrospective, population-based, cross-sectional study.

**Setting:** Ontario, Canada’s most populous province.

**Participants:** Patients who had received at least one family physician visit at one of 13 virtual walk-in clinics from April 1^st^ to December 31^st^, 2020. They were compared to Ontario residents who had any virtual family physician visit in the same time period.

**Main Outcome(s) and Measure(s):** Patient characteristics and 30-day post-visit healthcare utilization.

**Results:** Virtual walk-in patients (N=132,168) had fewer comorbidities and lower previous healthcare utilization than Ontarians with any virtual visit. Less than 0.1% of virtual walk-in visits were with a patient’s own family physician. Compared to Ontarians having any virtual family physician visit, virtual walk-in patients were significantly less likely to have a subsequent in-person visit with the same physician (0.2% vs. 11.0%, SMD = 0.48), more likely to have a subsequent virtual visit (30.3% vs. 21.9%, SMD = 0.19), and twice as likely to have an emergency department visit within 30 days (8.3% vs. 4.1%, SMD = 0.18), an effect that persisted after adjustment and across rurality groups: large urban (aOR 2.26; 95% CI 2.08-2.45), small urban (aOR 2.08; 95% CI 1.99-2.18), and rural (aOR 1.87; 95% CI 1.69-2.07).

**Conclusions and Relevance:** Compared to Ontarians attending any family physician virtual visit, virtual walk-in patients were less likely to have a subsequent in-person physician visit, but were more likely to visit the emergency department. Low continuity and the lack of physical examination may be contributing to increased emergency department utilization for virtual walk-in clinic patients.

## INTRODUCTION

Virtual walk-in clinics provide direct-to-consumer video, telephone, or text-based physician consultations, often through a mobile phone application. Prior to COVID-19, these clinics ostensibly helped meet a primary care need for people without a family physician, or those who couldn’t access their physician in a timely way, including those in rural settings.^1-3^ In Canada, Australia and the U.S., new COVID-19-related billing codes, intended to support virtual visits within existing primary care relationships, also drove a proliferation of virtual walk-in clinics.^1,4-8^ Many patients like virtual visits, particularly with their own physician, as they do not have to take time off work, arrange childcare, travel long distances or pay for parking.^9-15^

Despite these positive perceptions, there remain concerns about the quality of care provided through virtual visits in general, and in particular by virtual walk-in clinics.^16,17^ These offer an exclusively virtual experience, typically outside of existing primary care relationships.^17^ The lack of a physical exam has raised questions as to whether and how virtual encounters meet the standard of care for higher-acuity presentations.^18^ Exclusively virtual care clinics typically do not integrate with patients’ existing source of primary care, raising concerns about duplication and potential harm resulting from care discontinuity.^17^ Virtual visits may also exacerbate inequities in access resulting from language discordance, technological access or literacy.^16,19-21^ Fourth, the 24/7 access afforded by virtual walk-in clinics may prompt visits for transient, low-acuity medical symptoms that previously would not have occurred at all,^16^ raising total system costs – a phenomenon known as “supplier-induced demand.”^22^

Although previous studies have described the rapid expansion of virtual care, previous reports could not distinguish virtual walk-in clinic visits from all virtual primary care visits.^5,23^ Little is known about the physicians and patients who use exclusively virtual walk-in clinics. Our objectives were twofold: i) to describe the family physicians working in virtual walk-in clinics and compare these to the broader family physician pool and ii) to describe the characteristics and healthcare use of patients using virtual walk-in clinics compared to the general population, and a subset that received any virtual family physician visit.

## METHODS

### Study Design and Setting

We conducted a retrospective, population-based cross-sectional study of all Ontario residents, with a focus on family physicians and patients who participated in encounters at any of 13 selected virtual walk-in clinics in Ontario.

Ontario is Canada’s most populous province, with over 14.5 million residents. Provincial health insurance is provided to all permanent residents without premiums or co-payments, and covers emergency department visits, hospitalizations, and all medically necessary physician care. In Ontario, most primary care is provided by family physicians, and nearly 80% of the population is enrolled to a family physician working in a patient enrolment model.^24^

Prior to April 2020, use of an approved platform (the Ontario Telemedicine Network^15^) and a video (rather than telephone) visit were requirements to bill for a virtual visit in Ontario. After the onset of COVID-19, the Ministry of Health introduced several new temporary billing codes for synchronous virtual visits by video or phone with a value equivalent to that of in-person visits (Tables S1A-B). Since then, the majority of publicly-funded virtual visits are conducted by phone.^25,26^ Asynchronous visits, provided by email or text message, were not covered by provincial insurance.

This study was approved by the Women’s College Hospital Research Ethics Board (REB 2020-0095-E).

### Data Sources

Population-based health administrative datasets were linked using unique encoded identifiers and analyzed at ICES in Ontario, Canada. ICES is an independent, non-profit research institute whose legal status under Ontario’s health information privacy law allows it to collect and analyze health care and demographic data, without consent, for health system evaluation and improvement (see Table S2 for list of databases used).

In addition to ICES permanent datasets, we developed a non-comprehensive list of virtual walk-in clinics by searching business names. We obtained a list of group billing numbers and corresponding group names from the Ontario Ministry of Health. We used this list to identify all groups with “virtual” or “tele” in their name, Google-searched the identified names, and reviewed clinic websites to determine which provided exclusively virtual care, defined as virtual care without the possibility of an in-person office visit with a physician. In addition, we used Google to search the combined terms “Canada” or “Ontario” AND “virtual clinic” or “telemedicine”, thereby identifying several other groups for inclusion from our list of Ministry-provided group names and numbers, for a total of 20 virtual-only clinics. Within this list, we restricted to groups that had active billing claims during April 1^st^ 2019 to December 31^st^, 2020 (n = 13).

### Study Populations

#### Family physicians

We included all family physicians with a billing claim under one of our included virtual walk-in clinics from April 1^st^-December 31^st^ 2020. The comparison group was all family physicians with active billing from April 1^st^-December 31^st^, 2020.

#### Virtual walk-in clinic patients

We selected all patients who had received at least one family physician visit at one of the 13 included virtual walk-in clinics from April 1^st^-December 31^st^, 2020. The comparison group was all residents of Ontario with an active provincial health card as of April 1^st^, 2020, with any health care contact within the previous 8 years. For measures of healthcare utilization, we restricted the Ontario population to those who had at least one virtual family physician visit from April 1^st^ – December 31^st^, 2020.

#### Patient Characteristics and Healthcare Utilization

We reported the following patient characteristics: age, sex, neighborhood income quintile, urban/rural residence,^27^ and whether they were a recent provincial insurance registrant (within past 10 years), a proxy measure for recent immigration.^24^ We also examined the count of comorbidities using Johns Hopkins Aggregated Diagnosis Groups (ADGs; obtained from The Johns Hopkins ACG^®^ System Version 10) and prior healthcare utilization using ACG^®^ Resource Utilization Bands (RUBs) over the previous 2 years.^28,29^ We described patient enrolment status, enrolment model type, and continuity of care using Usual Provider Continuity (UPC)^30^ (see Table S3 for operational definitions of all variables).

For patients with more than one virtual clinic visit, we randomly selected a virtual clinic visit as the index date and excluded all others (Figure S1). For the Ontario population comparison, characteristics were anchored to April 1^st^, 2020, and for characteristics that required anchoring to an encounter, we randomly selected one virtual visit per patient and excluded all others.

We reported the frequencies of the top 10 most common medical diagnoses in each group. We also described whether virtual encounters were with a patient’s enrolling family physician, the encounter day of the week, as well as 30-day post visit healthcare utilization, including a repeat virtual visit, office physician visit, low-acuity emergency department visit (defined as Canadian Triage and Acuity Scale score of 4-5^31^), any emergency department visit, or urgent hospitalization.

#### Data Analyses

First, we plotted the weekly count of patients and physicians having encounters at virtual clinics, from April 2019 to December 2020. We then compared the characteristics of physicians who provided a virtual walk-in clinic visit in April-December 2020 to all family physicians with active billings in this period. We compared virtual walk-in clinic visit patients to the general Ontario population, and the subset of the Ontario population that received any virtual family physician visit. We also stratified healthcare utilization variables by patient rurality (large urban, small urban, and rural), since this is known to be associated with rates of emergency department use.^32^

To compare groups, we used standardized mean differences, and considered differences greater than 10% (0.1) to be meaningful.^33^ To examine the adjusted association between type of virtual visit with emergency department use in the subsequent 30 days, we used logistic regression with a generalized estimating equation (GEE) to account for clustering by the index virtual visit physician. Any patient who had received both types of virtual visit was removed from the “other virtual” visit group, such that each individual appeared only once. We stratified the regression by urban/rural category (large urban, small urban, and rural), and adjusted for patient age, sex, neighborhood income quintile, resource utilization band, and recent provincial insurance registrant status.

Analyses were executed in SAS software, version 9.4 (SAS Institute Inc., Cary, NC).

## RESULTS

### Virtual walk-in clinic volumes over time

From April 2019 to December 2020, the weekly volume of patients increased twofold from an average of 3,015 patients/week to an average 6,900 patients/week (Figure 1). The number of individual physicians providing virtual encounters each week increased sharply between March and May 2020, and by November 2020 was 2.5 times higher than in February 2020.

**Figure 1.**
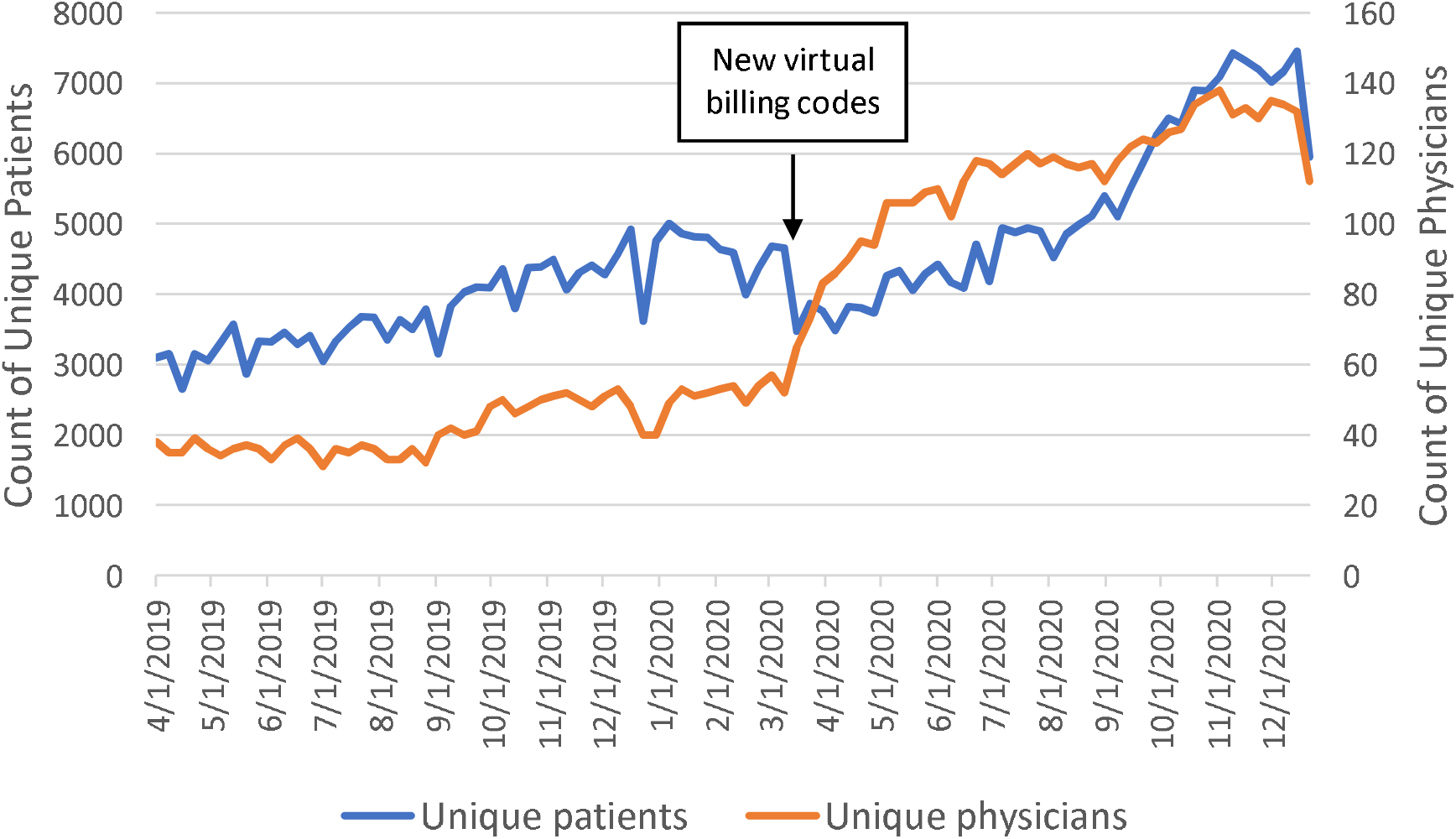
Weekly count of unique patients and unique physicians working for selected virtual walk-in clinics (N = 13) in Ontario, from the week of April 1^st^, 2019 to December 27^th^, 2020. New virtual billing codes introduced on March 14, 2020.

### Physicians working in virtual walk-in clinics

Compared to the overall Ontario population of family physicians with active billing between April 1^st^ and December 31^st^, 2020 (*n*=14,825; Table 1), virtual walk-in clinic physicians were younger, more likely to have graduated within the past 10 years and to practice in a large urban setting. They were also considerably more likely to work fee-for-service, outside of a patient enrolment model. There was no meaningful difference in the number of patients seen per day.

**Table 1.** Virtual Walk-In Clinic Physician Characteristics Compared to All Active Billing Family Physicians. Measured between April 1^st^, 2020 and December 31^st^, 2020.

| Physician Characteristics | Performed > 5 virtual walk-in clinic visits (N=242) | All family physicians with active billing (N=14,825) | Standardized mean difference* |
| --- | --- | --- | --- |
| **Physician age** |  |  |  |
| Mean ± SD | 40.3 ± 10.9 | 49.3 ± 14.0 | 0.72 |
| Median (IQR) | 37 (32-46) | 48 (38-59) | 0.75 |
| **Physician age group, n (%)** |  |  |  |
| 25 – 34 | 102 (42.1) | 2,471 (16.7) | 0.58 |
| 35 – 49 | 95 (39.3) | 5,285 (35.6) | 0.07 |
| 50 – 64 | 34 (14.0) | 4,843 (32.7) | 0.45 |
| 65+ | 11 (4.5) | 2,226 (15.0) | 0.36 |
| **Physician self-reported female gender, n (%)** | 119 (49.2) | 7,112 (48.0) | 0.02 |
| **Physician years since medical school graduation, n (%)** |  |  |  |
| 0 – 5 | 37 (15.3) | 764 (5.2) | 0.34 |
| 6 – 10 | 57 (23.6) | 2,280 (15.4) | 0.21 |
| 11 – 20 | 32 (13.2) | 2,939 (19.8) | 0.18 |
| 21 – 30 | 35 (14.5) | 3,027 (20.4) | 0.16 |
| 31+ | 20 (8.3) | 4,446 (30.0) | 0.57 |
| Missing | 61 (25.2) | 1,369 (9.2) | 0.43 |
| **Physician practice urban/rural location, n (%)** |  |  |  |
| Large urban | 202 (83.5) | 11,010 (74.3) | 0.23 |
| Small urban | 25 (10.3) | 2,303 (15.5) | 0.16 |
| Rural | 9 (3.7) | 1,043 (7.0) | 0.15 |
| Missing | 6 (2.5) | 469 (3.2) | 0.04 |
| **Physician primary care model, n (%)** |  |  |  |
| Enhanced fee-for-service | 58 (24.0) | 2,753 (18.6) | 0.13 |
| Capitation | 40 (16.5) | 4,665 (31.5) | 0.36 |
| Team-Based | 0 (0.0) | 1,824 (12.3) | 0.53 |
| Fee-For-Service (no enrolment) | 136 (56.2) | 4,926 (33.2) | 0.47 |
| Other | 8 (3.3) | 657 (4.4) | 0.06 |
| **Number of patients seen on a day doing virtual visits** |  |  |  |
| Median (IQR) | 12 (5 – 22) | 13 (7 – 22) | 0.05 |

Note. \*A standardized difference of at least 10% (0.1) was considered to indicate a meaningful difference.

### Patients attending virtual walk-in clinics

Compared to the overall Ontario population, patients who attended a virtual walk-in clinic visit were more likely to be adults between the ages of 18 and 44 years and less likely to be children (<18 years) or older adults (65-74 and 75+, Table 2). Virtual walk-in clinic patients were also more likely to be female and live in a small urban setting. The proportion of virtual walk-in clinic patients that were new registrants or that resided in low-income neighborhoods did not differ from the overall Ontario population.

**Table 2.**
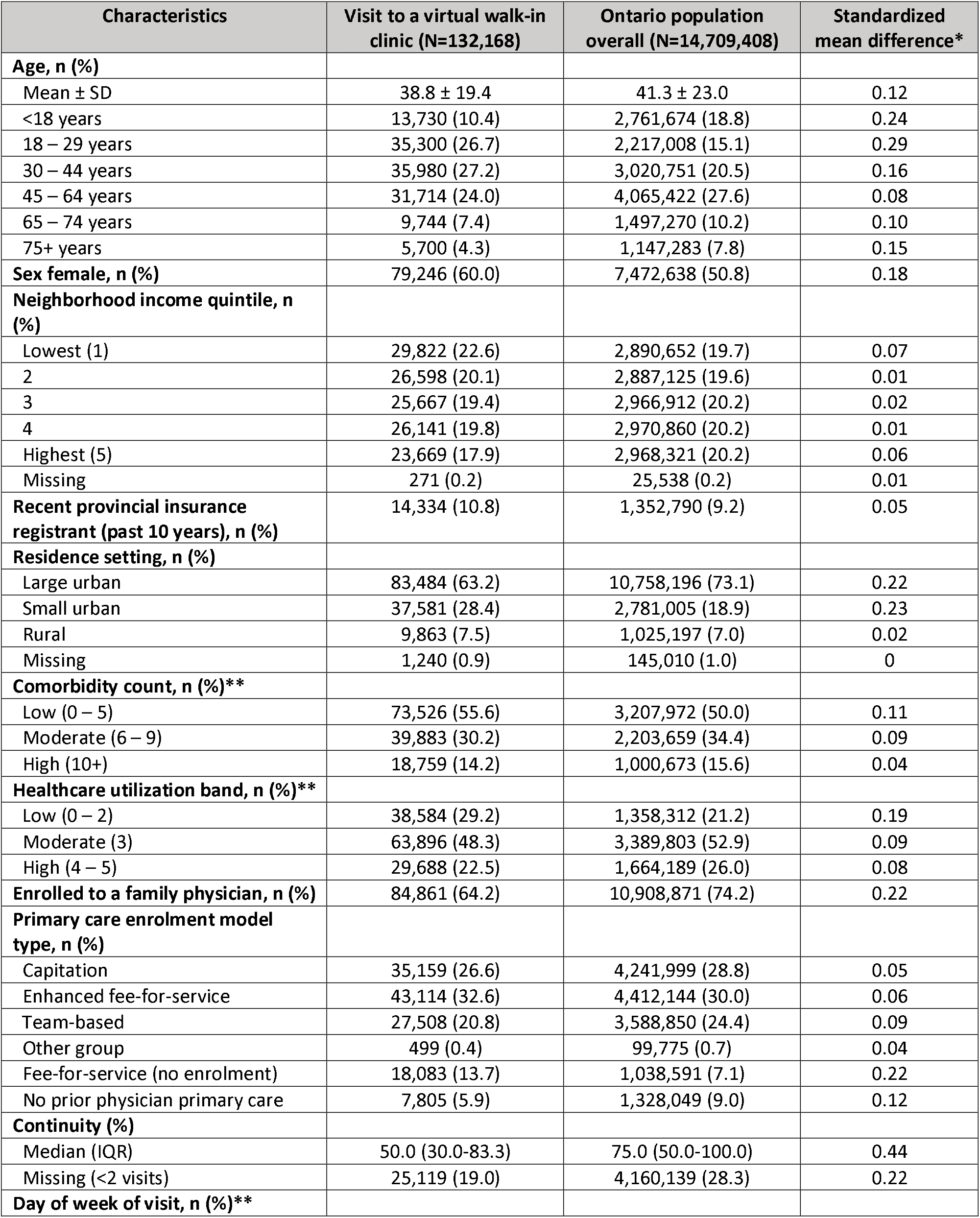
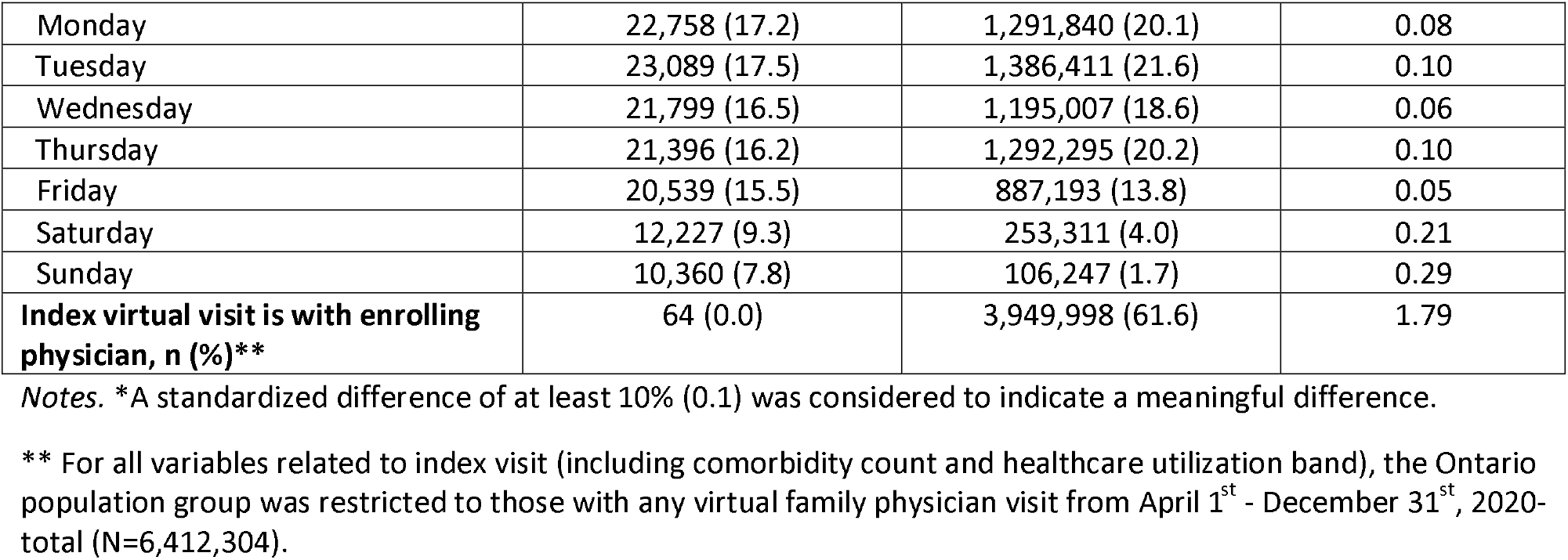
Patient Characteristics for Visits at Virtual Walk-In Clinics Compared to the Ontario Population with any Virtual Family Physician Visit. Measured between April 1^st^ and December 31^st^, 2020.

Virtual walk-in clinic patients were less likely to be enrolled to a family physician (64.2% vs. 74.2%, SMD = 0.22) and had lower continuity of primary care than the Ontario population (SMD = 0.44). Less than 0.1% of virtual walk-in visits were with the patient’s enrolling family physician.

Compared to all Ontarians who had any virtual family physician visit, virtual walk-in clinic patients had fewer comorbidities (55.6% vs. 50% were “low”, SMD = 0.11) and lower levels of previous healthcare utilization (29.2% vs. 21.2% were “low”, SMD = 0.19). They were also more likely to have their virtual visit on a Saturday or Sunday.

### Top 10 diagnoses at virtual clinic encounters

The top 10 diagnoses entered for virtual walk-in clinic visits were similar to those claimed for all Ontarians’ virtual family physician visits-5 out of 10 were present in both lists (Table 3). However, acute conditions (acute nasopharyngitis, coronavirus, cellulitis) occurred more commonly among virtual walk-in clinic visits, and chronic disease diagnoses (hypertension, diabetes mellitus, lipid disorders) were more common among all virtual family physician visits.

**Table 3.** Top 10 Diagnoses for Virtual Walk-In Clinic Visits and for the Ontario Population with any Virtual Family Physician Visit in 2020. Measured between April 1^st^, 2020 and December 31^st^, 2020.

| Virtual walk-in visit, n (%)<br>N=132,168 |  | Ontario population with virtual family physician<br>visit in 2020, n (%)<br>N=6,412,304 |  |
| --- | --- | --- | --- |
| Other ill-defined conditions | 13,837 (10.5) | Mental Health <sup>b</sup> | 488,468 (7.6) |
| Cystitis | 6,430 (4.8) | Other ill-defined conditions | 393,541 (6.1) |
| Mental health | 5,226 (4.0) | Essential, benign hypertension | 372,793 (5.8) |
| Acute nasopharyngitis, common cold | 4,838 (3.7) | Diabetes mellitus, including complications | 330,292 (5.2) |
| Coronavirus | 4,031 (3.1) | Musculoskeletal symptoms other than back pain <sup>c</sup> | 225,615 (3.5) |
| Gastrointestinal symptoms <sup>a</sup> | 3,598 (2.7) | Gastrointestinal symptoms <sup>a</sup> | 202,921 (3.2) |
| Other disorders of urinary tract | 3,518 (2.7) | Eczema, atopic dermatitis, neurodermatitis | 140,262 (2.2) |
| Essential, benign hypertension | 3,415 (2.6) | Disorders of lipid metabolism | 136,877 (2.1) |
| Cellulitis, abscess | 2,993 (2.3) | Acute nasopharyngitis, common cold | 135,912 (2.1) |
| Family planning, contraceptive advice, advice on sterilization or abortion | 2,778 (2.1) | Lumbar strain, lumbago, coccydynia, sciatica | 107,305 (1.7) |

<sup>a</sup> Gastrointestinal symptoms = “Anorexia, nausea and vomiting, heartburn, dysphagia, hiccup, hematemesis, jaundice, ascites, abdominal pain, melena, masses”
<sup>b</sup> Mental health = “Anxiety, neurosis, hysteria, neurasthenia, obsessive compulsive neurosis, reactive depression”
<sup>c</sup> Musculoskeletal symptoms other than back pain = “Leg cramps, leg pain, muscle pain, joint pain, arthralgia, joint swelling, masses”

### Healthcare utilization following first virtual visit

Patients of virtual walk-in clinics had more repeat virtual visits with any physician in the 30 days following their initial visit compared to the average Ontarian with a virtual family physician visit, (30.3% *vs*. 21.9%, SMD = 0.19; Table 4). They were also considerably less likely to have an in-person visit with the same physician (0.2% vs. 11.0%, SMD = 0.48), with any physician (11.7% vs. 15.3%, SMD = 0.11), or with their own physician (4.3% vs. 9.1%, SMD = 0.20). Patients of virtual walk-in clinics were twice as likely to have any emergency department visit (8.3% *vs*. 4.1%, SMD = 0.18), with similar results found when stratifying patient residence location by rurality (i.e., large urban, small urban, and rural; Table S4). Virtual walk-in patients were also twice as likely to have a low-acuity emergency department visit (2.7% vs. 1.1%, SMD = 0.12).

**Table 4.** Thirty-day Post-Visit Utilization for Virtual Walk-In Clinic Patients Compared to Ontario Population with a Virtual Family Physician Visit. Measured between April 1^st^, 2020 and December 31^st^, 2020.

| Measures of Utilization within 30 days following the virtual visit, n (%) | Visit to a virtual walk-in clinic (N=132,168) | Ontario population with virtual family physician visit in 2020 (N=6,412,304) | Standardized mean difference* |
| --- | --- | --- | --- |
| At least one repeated virtual visit with *any* physician | 40,030 (30.3) | 1,403,778 (21.9) | 0.19 |
| At least one in-person visit with *same* physician | 309 (0.2) | 704,759 (11.0) | 0.48 |
| At least one in-person visit with *own enrolling* physician | 5,633 (4.3) | 584,993 (9.1) | 0.20 |
| At least one in-person visit with *any* physician | 15,441 (11.7) | 980,556 (15.3) | 0.11 |
| At least one emergency department visit | 11,003 (8.3) | 262,509 (4.1) | 0.18 |
| At least one low-acuity emergency department visit | 3,517 (2.7) | 69,425 (1.1) | 0.12 |
| At least one urgent hospitalization | 1,178 (0.9) | 49,717 (0.8) | 0.01 |

*Note.* \*A standardized difference of at least 10% (0.1) was considered to indicate a meaningful difference.

After adjustment, those who received a virtual walk-in clinic visit remained more likely to have an emergency department visit within 30 days, in all three patient residence strata: large urban (adjusted odds ratio [OR] 2.26; 95% CI 2.08-2.45), small urban (adjusted OR 2.08; 95% CI 1.99-2.18), and rural locations (adjusted OR 1.87; 95% CI 1.69-2.07).

## DISCUSSION

We compared patient characteristics and outcomes from visits to 13 virtual walk-in clinics to all virtual family physician visits in the Ontario population. Virtual walk-in patients were younger, more likely to be female, and had lower continuity of care than the general population; they also had lower previous healthcare usage than Ontario residents with any virtual family physician visit. Compared to Ontarians attending any family physician virtual visit, virtual walk-in patients were more likely to have a repeat virtual visit and less likely to have a subsequent *in-person* physician visit in the subsequent 30 days. They were also significantly more likely to visit the emergency department, a finding that held true in big cities, small towns, and rural areas, even after adjustment for potential confounders.

Our findings highlight two areas of potential concern with virtual walk-in clinics. The first is the lack of continuity of patient/physician relationships, a limitation shared with regular walk-in clinics. This is almost certainly accompanied by a lack of informational continuity, as presently there are no incentives or even regulatory frameworks compelling a virtual (or non-virtual) walk-in physician to share information with a patient’s usual provider. Easy access to a family physician outside existing primary care relationships should be weighed against the risks of low-continuity care.^34-36^ Low-continuity care does not offer opportunities for longitudinal preventive care, and has been associated with more adverse events among patients with chronic conditions like diabetes.^37^

The second major concern highlighted by our findings is the potential downstream consequences of a care model that operates without the possibility of a physical examination. Patients who have a virtual visit with their own family physician have more options for in-person follow-up – we found that 11% of patients with any virtual physician visit had an in-person visit with their virtual visit physician within 30 days, compared to only 0.2% of the virtual walk-in patients. In the absence of a physical examination, physicians at virtual walk-in clinics may recommend that patients go to emergency departments to be assessed in person. Alternatively, our finding of higher rates of emergency department visits among virtual walk-in clinic users could reflect the downstream consequences of an incorrect or delayed diagnosis.

The absence of a physical examination also has the potential to negatively affect other dimensions of care quality,^16^ lead to more inappropriate prescriptions,^38-40^ testing, follow-up visits^41^, and referrals to consultants. Supplier-induced demand through attractive marketing campaigns, combined with increased downstream healthcare utilization, could increase overall healthcare costs.

Reports from the US, UK and Sweden have described virtual visit users as more likely to be healthy young adults^42,43^ with higher socioeconomic status.^13,21,44^ Although we similarly found that users were more likely to be young adults with lower levels of healthcare utilization, our findings do not suggest that publicly-funded virtual walk-in visits are disproportionately serving the affluent. We found that virtual walk-in clinic users were no more or less likely to be from low- or high-income neighborhoods than the general population using virtual care. Like others,^12,13^ we found that virtual walk-in doctors were younger, with fewer years in practice than the average family physician. They were also more likely to be fee-for-service physicians, who would have experienced a sharp drop in income early in the pandemic due to decreased in-person visit volumes^5^ as well as increased expenditures related to personal protective equipment and enhanced cleaning. Without the income stability offered by capitation payments, fee-for-service physicians likely turned to other revenue sources, including virtual walk-in clinics.

Developing a policy landscape that favours an optimal and efficient use of virtual visits is an urgent priority for health insurers.^1,45,46^ In 2021, the Ontario Ministry of Health added new virtual visit codes to the “outside use” list, which financially penalises physicians funded through capitation models each time their patients see other family physicians.^47^ Policymakers seeking to direct virtual visit funding to existing primary care relationships could also consider significantly reducing the value of virtual visit codes when used by providers without a physical office location, or without a pre-existing primary care roster. Another option is for physician regulatory bodies to mandate that physicians offering virtual visits also offer in-person appointments, as was recently done by the College of Physicians and Surgeons of Manitoba.^18^

Our study has several limitations. First, we could only capture a subset of all virtual clinic encounters. There are likely many more physicians and patients participating in virtual walk-in clinic care, however because these are either not linked to a group billing number, or are privately paid,^4^ we had no way of identifying them for study inclusion. Second, we could not distinguish video from phone visits, as these were claimed using the same billing code. We further could not capture text (“e-visits”) or email consultations, as these are ineligible for coverage by provincial insurance and would be all privately paid. Third, we exclusively focused on family physicians, as these are the most common providers of primary care in Ontario, but did not assess visits to pediatricians or psychiatrists. Finally, our findings are most generalizable to other settings with publicly-funded virtual walk-in visits.

## Conclusion

The number of Ontario patients and family physicians participating in a sample of virtual walk-in clinics rose rapidly subsequent to COVID-19-related physician fee schedule changes. Our findings suggest that these low-continuity visits without a physical examination were associated with increased emergency department utilization. To ensure virtual walk-in clinics contribute positively to health outcomes and health system efficiency, policymakers should prioritize regulations and billing changes that ensure the integration of virtual/in-person visits and promote continuity of care.

## Supporting information

STROBE checklist

## Data Availability

The dataset from this study is held securely in coded form at ICES. While legal data sharing agreements between ICES and data providers (e.g., healthcare organizations and government) prohibit ICES from making the dataset publicly available, access may be granted to those who meet pre-specified criteria for confidential access, available at www.ices.on.ca/DAS. The full dataset creation plan and underlying analytic code are available from the authors upon request, understanding that the computer programs may rely upon coding templates or macros that are unique to ICES and are therefore either inaccessible or may require modification

## Acknowledgements

We would like to thank our patient partners Cherryl Bird, Krysta Nesbitt, Jerome Johnson and Patrick Roncal, for their contributions to discussions about our study findings. We also thank Alexander Kopp from ICES for his guidance in data analysis and methodology.

This project is supported by a Canadian Institutes of Health Research (CIHR) project grant (#175285). Lauren Lapointe-Shaw is supported by the University of Toronto Department of Medicine, the Toronto General Hospital Research Institute, the Women’s College Institute for Health System Solutions and Virtual Care (WIHV), and the Peter Gilgan Centre for Women’s Cancers at Women’s College Hospital, in partnership with the Canadian Cancer Society. Tara Kiran is the Fidani Chair of Improvement and Innovation at the University of Toronto. She is supported as a Clinician Scientist by the Department of Family and Community Medicine at St. Michael’s Hospital and the University of Toronto. Noah M. Ivers is supported by a Canada Research Chair in Implementation of Evidence-based Practice and a Clinician Scholar award from the Department of Family and Community Medicine at Women’s College Hospital and the University of Toronto.

This study was supported by ICES, which is funded by an annual grant from the Ontario Ministry of Health (MOH) and the Ministry of Long-Term Care (MLTC). Parts of this material are based on data and information compiled and provided by: MOH, Canadian Institute for Health Information (CIHI), and Cancer Care Ontario (CCO). The analyses, conclusions, opinions and statements expressed herein are solely those of the authors and do not reflect those of the funding or data sources; no endorsement is intended or should be inferred.

