## Appendix for "Characteristics and healthcare use of patients attending virtual walk-in clinics: a cross-sectional analysis"

**Table S1A.** Temporary Ontario Health Insurance Plan Codes for Virtual Visits (Telephone and Video) and their In-Person Equivalents as of March 14, 2020

**Table S1B.** Ontario Telemedicine Network Virtual Care Program Billing Amendments to Enable Direct-to-Patient Video Visits, effective November 15, 2019 to March 31, 2020

**Table S2.** ICES Data Sources

**Table S3.** Operational Definitions of all Variables

**Table S4.** Post-Visit Utilization for Virtual Walk-In Clinic Patients Compared to Ontario Population with a Virtual FP/GP Visit Stratified by Large Urban, Small Urban, and Rural Patient Residence

**Figure S1.** Cohort Inclusion/Exclusion Flowcharts

**Table S1A. Temporary Ontario Health Insurance Plan Codes for Virtual Visits (Telephone and Video) and their In-Person Equivalents as of March 14, 2020.**

Available: <https://www.health.gov.on.ca/en/pro/programs/ohip/bulletins/4000/bul4745.pdf>
<https://www.health.gov.on.ca/en/pro/programs/ohip/sob/physserv/sob_master.pdf>

| **Code**  **(Virtual/In-Person Equivalent)** | **Description** | **Fee**  **(Virtual/In-Person Equivalent)** |
| --- | --- | --- |
| K080/A001 | Minor assessment of a patient, or advice or to a patient’s representative regarding health maintenance, diagnosis, treatment and/or prognosis. | $23.75/$23.75 |
| K081/A007 | Intermediate assessment of a patient, or advice or information to a patient’s representative regarding health maintenance, diagnosis, treatment and/or prognosis if the service lasts a minimum of 10 minutes. | $36.85/$36.85 |
| K082/K005 | Psychotherapy, psychiatric, or primary mental health care, counselling, or interview per unit (half hour or major part thereof). | $67.75/$67.75 |

**Table S1B. Ontario Telemedicine Network Virtual Care Program Billing Amendments to Enable Direct-to-Patient Video Visits, effective November 15, 2019 to March 31, 2020.**

Available: <https://www.health.gov.on.ca/en/pro/programs/ohip/bulletins/4000/bul4731.pdf>

| **Code** | **Description** | **Fee** |
| --- | --- | --- |
| B099A | Direct-to-patient video visit tracking code | $0.00 |
| B100A | Hosted video first telemedicine patient encounter premium | $35.00 |
| B200A | Hosted video subsequent telemedicine patient encounter premium | $15.00 |
| B101A | Hosted video first cancelled/missed telemedicine patient encounter premium | $35.00 |
| B201A | Hosted video subsequent cancelled/missed telemedicine patient encounter premium | $15.00 |
| B102A | Hosted video first technical difficulties abandoned patient encounter premium | $35.00 |
| B202A | Hosted video subsequent technical difficulties abandoned patient encounter premium | $15.00 |

*Note.* Direct-to-patient video visit codes are only eligible for delivery by specialist, GP focused-practice designated physicians, and primary care physicians who are in a PEM and are delivering care to a rostered patient.

**Table S2. ICES Data Sources**

| **Database name** | **Description** |
| --- | --- |
| **Client Agency Program Enrolment Database (CAPE)** | Links physicians to their enrolled patients under several patient enrolment models of clinical practice. These funding models include enhanced fee for service, non-team capitation, and team-based capitation. (1) |
| **Corporate Provider Database (CPBD)** | Information on providers (physicians, nurses, etc.) and groups (primary care, hospitals, etc.) eligible to receive payment from OHIP, such as physician demographics, training, and practice location. (2) |
| **Discharge Abstract Database (DAD)** | Information on all admissions (excluding designated mental health beds) to acute care hospitals in Ontario. This includes dates of admission as well as diagnostic and procedural codes. Overall, diagnostic codes were found to be 82% sensitive for primary diagnosis when verified against chart abstraction. (3) |
| **ICES Physician Database (IPDB)** | Contains yearly (fiscal) information about all physicians in Ontario. It is used to describe physician characteristics, such as sex, speciality, location, and measures of physician activity (billings, workload, types of services provided). (4) |
| **National Ambulatory Care Reporting System (NACRS)** | Includes information for all emergency department visits since 2000. A re-abstraction study of diagnostic codes found 85% agreement for the main presenting problem. (5) |
| **Ontario Health Insurance Plan (OHIP)** | Contains information on all billing claims submitted by Ontario physicians (consultations and procedures). Fee for service is the primary method of remuneration for 95% of specialist physicians and 50% of primary care physicians in Ontario. However, physicians practicing in non fee-for-service models submit shadow billings to OHIP, which appear as billing claims with a payment value of $0. (6) |
| **Primary Care Population (PCPOP)** | Population-level dataset that includes all people in Ontario who are deemed alive and eligible at a given point in time. All indicators are as of the index date, with various look back periods. The dataset contains information on demographics, primary care rostering, patient enrollment (traditional and virtual), chronic disease flags, emergency department visits, hospital readmissions, ambulatory care sensitive conditions, screening, diabetic care, count of comorbidities, healthcare utilization, specialist visits, and continuity of care. (7) |
| **Registered Persons Database (RPDB)** | Contains demographic information about anyone who has ever received an Ontario health card number, i.e. all Ontarians alive at any time since 1990 (over 16 million records). (8) |

**Table S3. Operational Definitions of all Variables.**

| **Patient-Level Variables** | **Data Source** | **Definition** |
| --- | --- | --- |
| Age | PCPOP | Categorized as:  <18 years (children)  18-39 years (young adult)  40-64 years (middle aged)  65-79 years (younger seniors)  80+ years (older seniors) |
| Sex | PCPOP | Male Female |
| Neighborhood income quintile | PCPOP | Nearest census-based income quintile based on postal code (based on 2016 census).(9) |
| Recent OHIP registrant | PCPOP | New OHIP registrant within the previous 10 years, a proxy for immigration. |
| Residence setting | PCPOP | Postal code converted to RIO score (10):  0-9: Large urban  10-40: Small Urban  40+: Rural  Missing |
| Comorbidity count | DAD, NACRS, OHIP | Count of ACG System Aggregated Diagnosis Groups (ADGs, per the Johns Hopkins ACG® System Version 10, in 2 years prior to April 1^st^, 2019 (for pre-COVID group) or April 1^st^, 2020 (for post-COVID group).(11) Categorized as:  Low: 0-5  Moderate: 6-9  High: 10+ |
| Healthcare utilization band | DAD, NACRS, OHIP | Using Resource Utilization Bands (RUBs), per the Johns Hopkins ACG® System Version 10, in 2 years prior to April 1^st^, 2019 (for pre-COVID group) or April 1^st^, 2020 (for post-COVID group).(11) Categorized as:  Low: (0-2)  Moderate: (3)  High: (4-5) |
| Enrollment to a primary care physician | PCPOP | Whether patient is enrolled (R_TYPE = R) |
| Patient primary care enrolment model type | PCPOP | Variable PROGTYPE 3 from PCPOP, grouped into:  Capitation  Team-Based (Family Health Team)  Enhanced Fee-For-Service (Comprehensive care model or family health group)  Other (all other models)  Fee-For-Service (no enrolment)  No group  No primary care |
| Continuity | PCPOP | Proportion of visits to the patients’ primary care provider out of all visits, over previous 2 years. Code those with <2 visits as missing. |
| Day of the week of visit | OHIP | Categorized into:  Monday  Tuesday  Wednesday  Thursday  Friday  Saturday  Sunday |
| **Physician-Level Variables** | **Data Source** | **Definition** |
| Physician age | CPDB | Mean physician age |
| Physician age group | CPDB | Physician age based on birth year (BYEAR) categorized into:  25-34  35-49  50-64  65+ |
| Self-reported physician gender | CPDB | Male or female. |
| Physician years since graduation | IPDB | Years since graduation (GRADYEAR) in the following categories:  0-5  6-10  11-20  21-30  31+ |
| Physician practice urban/rural location | CPDB | Categorized based on RIO score of physician practice.  0-9: large urban  10-39: small urban  40+: rural |
| Physician primary care model | CAPE | Capitation  Team-Based (Family Health Team)  Enhanced Fee-For-Service (Comprehensive care model or family health group)  Other (all other models)  Fee-For-Service |
| Number of patients seen on a day doing virtual visits | OHIP | Median number of patients seen on a typical day doing virtual visits. |
| **Measures of Utilization within 30 days following the virtual visit** | **Data Source** | **Definition** |
| At least one repeated virtual visit with any physician | OHIP | Proportion of virtual visits with a subsequent virtual visit with any FP/GP physician following 30 days of the index virtual visit. |
| At least one in-person visit with same physician | OHIP | Proportion of virtual visits with a subsequent in-person visit (office or home) with the same physician within 30 days of the index visit. |
| At least one in-person visit with any physician | OHIP | Proportion of in-person visits (office or home) following a virtual visit within 30 days following the index virtual visit. |
| At least one in-person visit with own enrolling physician | PCPOP  OHIP | Proportion of in-person visits (office or home) with the patient’s own rostered primary care provider within 30 days the index virtual visit. |
| At least one emergency department visit | NACRS | Code as true if visit to the emergency department within 30 days of virtual visit. Start count on the day of the index virtual visit date. |
| At least one emergency department visit with CTAS 4/5 | NACRS | Code as true if visit to the emergency department within 30 days of the index virtual visit had a CTAS of 4 or 5. Start the count on the day of the index virtual visit date. |
| At least one urgent hospitalization | DAD | Code as true if urgent admission (ADMCAT= ”U”) to an acute care hospital within 30 days of virtual visit. Start the count on the day of the index virtual visit date. |

**Table S4. Post-Visit Utilization for Virtual Walk-In Clinic Patients Compared to Ontario Population with a Virtual Family Physician Visit Stratified by Large Urban, Small Urban, and Rural Patient Residence.** Measured from April 1^st^ to December 31^st^, 2020.

| **Measures of Utilization** | **Visit to a virtual walk-in clinic** | | | **Ontario population with virtual family physician visit in 2020** | | | **Standardized mean difference*** | | |
| --- | --- | --- | --- | --- | --- | --- | --- | --- | --- |
|  | **Large Urban**  **(N=83,484)** | **Small Urban**  **(N=37,581)** | **Rural**  **(N=9,863)** | **Large Urban**  **(N=4,899,462)** | **Small Urban**  **(N=1,118,636)** | **Rural**  **(N=354,926)** | **Large Urban** | **Small Urban** | **Rural** |
| **At least one repeated virtual visit with any physician within the next 30 days, n (%)** | 26,730 (32.0) | 10,503 (27.9) | 2,498 (25.3) | 1,137,217 (23.2) | 202,224 (18.1) | 57,110 (16.1) | 0.20 | 0.24 | 0.23 |
| **At least one in-person visit with same physician within the next 30 days, n (%)** | 208 (0.2) | 70 (0.2) | 29 (0.3) | 551,929 (11.3) | 113,705 (10.2) | 35,299 (9.9) | 0.49 | 0.46 | 0.45 |
| **At least one in-person visit with any physician within the next 30 days, n (%)** | 10,869 (13.0) | 3,708 (9.9) | 764 (7.7) | 775,069 (15.8) | 153,718 (13.7) | 46,236 (13.0) | 0.08 | 0.12 | 0.17 |
| **At least one in-person visit with own enrolling physician within the next 30 days, n (%)** | 3,629 (4.3) | 1,628 (4.3) | 342 (3.5) | 451,543 (9.2) | 100,147 (9.0) | 30,456 (8.6) | 0.19 | 0.19 | 0.22 |
| **At least one emergency department visit within the next 30 days, n (%)** | 6,318 (7.6) | 3,507 (9.3) | 1,043 (10.6) | 181,880 (3.7) | 55,209 (4.9) | 22,749 (6.4) | 0.17 | 0.17 | 0.15 |
| **At least one urgent hospitalization within the next 30 days, n (%)** | 766 (0.9) | 321 (0.9) | 74 (0.8) | 35,235 (0.7) | 10,080 (0.9) | 3,916 (1.1) | 0.02 | 0.01 | 0.04 |

*Note.* *A standardized difference of at least 10% (0.1) was considered to indicate a meaningful difference.

**Figure S1. Cohort inclusion/exclusion flow charts**

Ontario population as of April 1^st^, 2020

(PCPOP dataset)

N = 14,709,408 individuals

Ontario population with a virtual family physician visit

April 1^st^–December 31^st^ 2020.

N = 6,412,304 individuals

No virtual family physician visit between April 1^st^-December 31^st^, 2020

N = 8,297,104 individuals

Encounters in virtual walk-in clinics April 1^st^-December 31^st^, 2020

n = 224,211 encounters

Randomly select one virtual visit per patient

n = 132,168 individuals

4. ICES. Physician Database (IPDB). In ICES Data Dictionary; 2018.

5. Canadian Institute for Health Information. CIHI Data Quality Study of Ontario Emergency Department Visits for Fiscal Year 2004–2005—Executive Summary. Ottawa; 2007.

6. ICES. Ontario Health Insurance Plan (OHIP). In ICES Data Dictionary; 2021.

7. ICES. Primary Care Population (PCPOP). In ICES Data Dictionary; 2021.

8. ICES. Population and Demographic Data (RPDB). In ICES Data Dictionary; 2021.

9. Alter DA, Naylor CD, Austin P, Tu JV. Effects of Socioeconomic Status on Access to Invasive Cardiac Procedures and on Mortality after Acute Myocardial Infarction. New England Journal of Medicine 1999;341:1359-1367.

10. Kralj B. Measuring Rurality - RIO2008_BASIC: Methodology and Results In: OMA Economics Department, editor.; 2009.

11. Johns Hopkins University. Johns Hopkins ACG Case-Mix Adjustment System.
